# Treatment with an Anti-CK2 Synthetic Peptide Improves Clinical Response in Covid-19 Patients with Pneumonia. A Randomized and Controlled Clinical Trial

**DOI:** 10.1101/2020.09.03.20187112

**Authors:** Leticia R. Cruz, Idania Baladrón, Aliusha Rittoles, Pablo A. Díaz, Carmen Valenzuela, Raúl Santana, Maria M. Vázquez, Ariadna García, Deyli Chacón, Delvin Thompson, Gustavo Perera, Ariel González, Rafael Reyes, Loida Torres, Jesus Pérez, Yania Valido, Ralysmay Rodriguez, Dania M. Vázquez, Mauro Rosales, Ailyn C. Ramón, George V. Pérez, Gerardo Guillén, Verena Muzio, Yasser Perera, Silvio E. Perea, for the ATENEA-Co-300 group

## Abstract

**Purpose:** The instrumental role of CK2 in the SARS-Cov2 infection has pointed out this protein kinase as a promising therapeutic target in Covid-19. Anti-SARS-Cov2 activity has been reported by CK2 inhibitors *in vitro;* however, any anti-CK2 clinical approach has been investigated in Covid-19. This exploratory trial aimed to explore safety and putative clinical benefit of CIGB-325, an anti-CK2 peptide previously assessed in cancer.

**Methods:** A monocentric, parallel group design, therapeutic exploratory trial of intravenous CIGB-325 in adults hospitalized with Covid-19 was performed. Twenty patients were randomly assigned to receive CIGB-325 (2.5 mg/kg/day during 5-consecutive days) plus standard-of-care (10 patients) or standard-of-care (10 patients). Adverse events were classified by the WHO Adverse Reaction Terminology. Parametric and non-parametric statistical analyses were performed according to the type of variable. Considering the small sample size, differences between groups were estimated by Bayesian analysis.

**Findings:** CIGB-325 induced transient mild and/or moderate adverse events like pruritus, flushing and rash in some patients. Both therapeutic regimens were similar respect to SARS-Cov2 clearance in nasopharynx swabs over the time. However, CIGB-325 significantly reduced the median number of pulmonary lesions (9.5 to 5.5, p = 0.042) at day 7 and proportion of patients with such effect was also higher according to Bayesian analysis (pDif > 0; 0.951). Additionally, CIGB-325 significantly reduced the CPK (p = 0.007) and LDH (p = 0.028) plasma levels at day 7.

**Implications:** Our preliminary findings suggest that this anti-CK2 clinical approach could be combined with standard-of-care in Covid-19 thus warranting larger studies.

## INTRODUCTION

Severe acute respiratory syndrome coronavirus 2 (SARS-CoV-2) has spread worldwide and has infected nearly 20 million people [1, 2]. Many infected people are asymptomatic or experience mild symptoms and recover without medical intervention [3, 4]. However, older people and those with comorbid hypertension, diabetes, obesity, and heart disease are at higher risk of life-threatening illness [5, 6]. Therefore, development of an effective antiviral drug for COVID-19 is a global health priority. Along with the development of new antiviral drugs, repurposing existing drugs for COVID-19 treatment is also accelerated [7]. Some antiviral drugs have shown high efficacy against SARS-CoV-2 both *in vitro* [8] and *in vivo* models [9, 10]. A number of clinical studies such as compassionate use programs and clinical trials have been conducted to test the efficacy of FDA-approved drugs, such as lopinavir and ritonavir, chloroquine, favipiravir, and remdesivir (RDV) [11-14]. More recently, a double-blind, randomized, placebocontrolled trial of intravenous RDV in adults hospitalized with Covid-19 and lower respiratory tract involvement had a median recovery time of 11 days (95% confidence interval [CI], 9 to 12), as compared with 15 days (95% CI, 13 to 19) in those who received placebo (rate ratio for recovery, 1.32; 95% CI, 1.12 to 1.55; P < 0.001) [15].

At present, the main antiviral strategies currently employed against SARS-CoV-2 can broadly be divided into two types: strategies directly targeting the virus and strategies indirectly targeting the virus via host modulation [16]. In both strategies there are already-approved drugs and experimental candidates in Clinical trials [16]. Likewise, discovery of novel cellular targets for SARS-Cov2 coronavirus has led to novel putative clinical strategies which merit going into clinical research in Covid-19.

CK2 is a constitutively active Ser/Thr protein kinase deregulated in cancer and other pathologies, responsible for about the 20% of the human phosphoproteome [17]. In infectious diseases, CK2 phosphorylates and modulates the function of viral proteins from Hepatitis C Virus (HCV), Vesicular stomatitis virus (VCV), Human Immunodeficiency Virus (HIV), Human Papilloma Virus (HPV), and Herpes simplex-1 (HSV-1) [18]. Therefore, it is expected that pharmacological intervention with CK2 may impact on in any step of viral life cycle. Recently, CK2 has been found to be directly targeted by the SARS-Cov2 nucleocapsid protein and they both co-localize along the filopodia protrusions that promote virus egress and rapid cell-to-cell spread across epithelial mono-layers of infected cells [19]. Additionally, inhibition of CK2 in those experiments demonstrated the instrumental role of this protein kinase for SARS-Cov2 infection *in vitro* [19].

Considering the scientific rationality of inhibiting CK2 in SARS-Cov2 infection and taking advance of CIGB-325 (formerly CIGB-300) as an anti-CK2 synthetic peptide [20, 21] previously already assessed in different Phase I-II in cancer patients, we investigated the putative clinical benefit of this peptide in Covid-19 patients. Particularly, safety and tolerability of intravenous delivery of CIGB-325 has been previously assessed from 0.2 to 12.8 mg/kg [22, 21] as well as the consecutive-5 day regimen of administration.

Based on the FDA recommendations for Covid-19 developing drugs, we performed a small and controlled clinical study [24]. In this exploratory study (proof-of-concept), we administered intravenous CIGB-325 at 2.5 mg/kg along with standard-of-care for treating SARS-Cov2 positive patients in Cuba which is based on alpha 2b-IFN plus kaletra/hydroxyquinoline. Preliminary data indicated that combination of CIGB-325 with standard-of-care improved chest-CT outcomes in Covid-19 patients with pneumonia at day 7. Other signs of clinical benefit for this therapeutic regimen were also registered in our study. This is the first clinical study where an anti-CK2 approach is explored in Covid-19 disease and the preliminary data provided here could warrant larger studies.

## PATIENTS AND METHODS

### Participants

From June 1 to June 16, 2020, twenty patients confirmed as SARS-CoV-2 positive by real-time transcriptase polymerase chain reaction (RT-PCR) were enrolled in a monocentric parallel group design in therapeutic exploratory trial at the “Luis Diaz Soto” Hospital in Havana, Cuba (https://rpcec.sld.cu/trials/RPCEC00000317-En, Code: IG/CIGB300I/CV/2001, ATENEA-Co-300 trial). CIGB-300 code used for cancer treatment was substituted by CIGB-325 just after the online registration of this clinical trial; therefore, it appears labeled with the former code. The Ethics Committee for Clinical Research in the hospital and Cuban Regulatory Agency (CECMED) approved the trial. The study complied with the Good Clinical Practices and the precepts established in the Declaration of the Helsinki World Medical Association. All patients met the inclusion criteria described in the protocol and signed the informed consent.

### Laboratory examination

Laboratory results included blood routine, leucocyte subsets and blood biochemical parameters. Serum levels of CPK, LDH and RCP were determined by a specific Roche system (Roche-cobas-C311).

### SARS-Cov2 viral dynamic in nasopharyngeal swabs

Nasopharyngeal swabs were obtained from patients at days 0, 3, 7, and 14. Viral RNA was detected by RT-PCR amplification by using specific SARS-Cov2 primers. Presence of SARS-Cov2 in swabs was followed-up at indicated times and compared among both groups.

### Clinical response

Clinical status was classified as asymptomatic, mild, moderate and severe Covid-19 disease according to the NIH guide “Coronavirus Disease 2019 (COVID-19) Treatment guidelines”. https://www.covid19treatmentguidelines.nih.gov. The chest-CT analysis was performed considering number of pulmonary lesions, lesion’s extent and common Covid-19 typical abnormalities like consolidation, glass-round opacity and mix pattern.

### Safety

Pretreatment evaluation included a detailed history and physical examination. In addition, hematological counts, blood chemistry, coagulation, radiography and chest-CT studies were performed. Systemic toxicity was evaluated daily after each CIGB-325 administration. Severity of adverse events was classified by the WHO Adverse Reaction Terminology. Causal relationship was classified as very probable (definitive), probable, possible or remote (doubtfull).

### Statistical Analysis

For chest-CT variables, the analysis was performed in Per Protocol population defined as patients who completed the originally allocated and patients that have TAC at the beginning and at the end of treatment. Continuous variables were expressed as mean and standard deviations or median and interquartile ranges (depending of the assumption of normal distribution). Student-t test or Mann-Whitney U test were applied to continuous variables, and chi-square or Fisher’s exact test were used for categorical variables. For the variation in time in each group, Wilcoxon-rank test or Student-t test for dependent variables were used. Type 1 error of 0.05 was specified. Considering the small sample size, the difference between groups (for clinical and chest-CT evaluations, proportion of patients with reduction of score and number of lesions) was estimated from the Bayesian point of view, performing 10,000 simulations and specifying non-informative prior distributions. The analysis was performed using SPSS 25.0 software and EPIDAT 3.1.

## RESULTS

### Baseline characteristics of Covid-19 patients

Between June 1, 2020, and June 16, 2020, twenty SARS-Cov2 positive patients underwent randomization in the hospital “Luis Diaz Soto” in Havana, Cuba. A flowchart of the study procedure is presented in Fig. 1. Ten were assigned to receive intravenous CIGB-325 + standard-of-care (Group I) and 10 to receive standard-of-care as control (Group II). To diminish odds of severity, the Cuban National Program for managing Covid-19 patients applies standard-of-care just upon confirmation of nasopharyngeal SARS-Cov2 diagnosis irrespectively of having symptoms or not. The median number of days between SARS-Cov2 diagnosis, hospitalization and treatment initiation was 2 days. All the patients completed the treatment as planned except one patient from Group I who experienced some concomitant moderate histaminergic adverse events during the first CIGB-325 intravenous delivery and dose was decreased to 1.6 mg/kg for remaining days as indicated in the clinical protocol.

**Figure 1:**
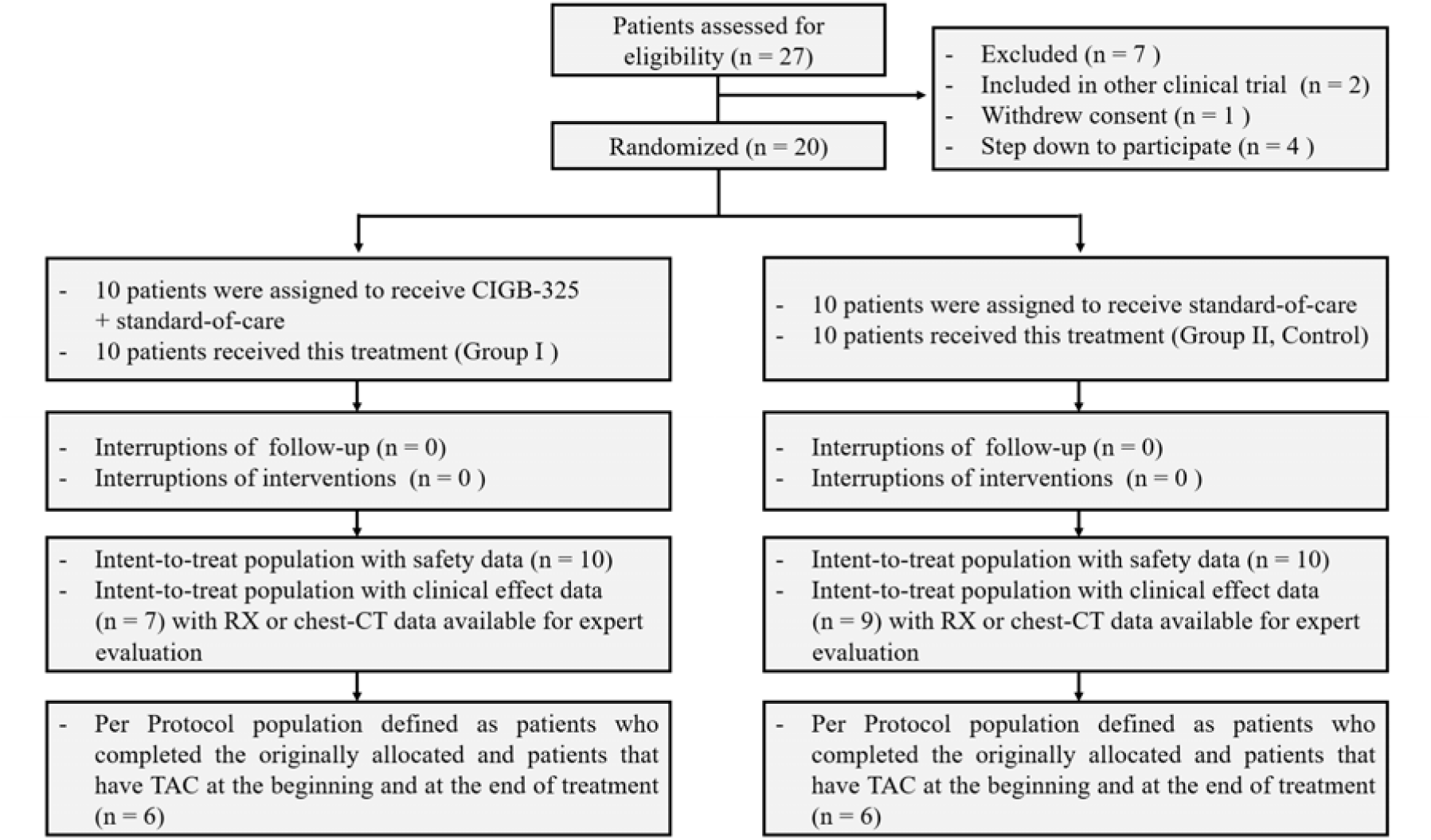
Randomization and enrollment.

Demographic characterizes among both groups were similar except the age. Fourteen patients (70%) were asymptomatic at the enrollment and 9 (45%) at the starting day of treatment. Overall, 25% of patients had hypertension, 25% had obesity and none had type 2 diabetes (Supp.1).

Data from baseline chest-CT analysis showed that 80% of patients in the CIGB-325 group had positive chest-CT according to presence of ground-glass opacity, consolidation, mix pattern and affectation of more than three pulmonary lobules. The mix pattern was the most predominant lesion. In the control group, 50% of patients had positive chest-CT with presence of consolidation, mixed pattern and affectation of less than two pulmonary lobules.

Concerning hematological and biochemical baseline parameters in all patients, abnormal levels were observed in hemoglobin (25%), platelets (15%), neutrophils (50%), lymphocytes (40%), aspartate aminotransferase (ASAT) (20%), alanine aminotransferase (ALAT)(40%), ferritin (35%), creatinine (20%), and glycemia (30%). Other parameters were in the normal range.

### Primary and exploratory efficacy end points

The dynamic conversion from positive to negative of SARS-Cov2 RT-PCR results was analyzed in both groups of treatments at 0, 3, 7, and 14 days. No significant differences were observed in the median time, 11 days ± 8.0 for CIGB-325 plus standard care and 12 days ± 6 for standard care alone (p = 0.614). Thus, time to SARS-Cov2 viral clearance in the nasopharynx swabs behaved similarly for both treatments over the time.

The clinical status was classified in categories of asymptomatic, mild, moderate or severe disease and it was followed up until ending treatment and thereafter for both groups. In the intent-to-treat population of CIGB-325 group at day 6 there were 2/10 patients that remained asymptomatic since initiation of treatment, 2/10 changed from severe to moderate, 1/10 from moderate to asymptomatic, 1/10 from mild to asymptomatic and 4/10 did not change their clinical status during the treatment. Otherwise, in the control 6/10 patients remained asymptomatic since initiation of treatment, 1/10 changed from mild to asymptomatic and 3/10 did not change their clinical status during the treatment. Additionally, chest-CT analysis was performed to investigate the effect of CIGB-325 over the Covid-19 pulmonary lesions. For that purpose, both number and lesion’s extent at day 0 and after treatment (day 7) were compared just in those patients analyzed per protocol (Table 1). Importantly, CIGB-325 treatment significantly reduced the median number of pulmonary lesions from 9.5 ± 10 (day 0) to 5.5 ± 10 (day 7) (p = 0.042). Conversely, no substantial change was observed in the control group. Proportion of patients with reduction of pulmonary lesions was higher in the CIGB-325 group compared with control according to the Bayesian analysis (pDif > 0; 0.951) (Table 1).

**Table 1:**
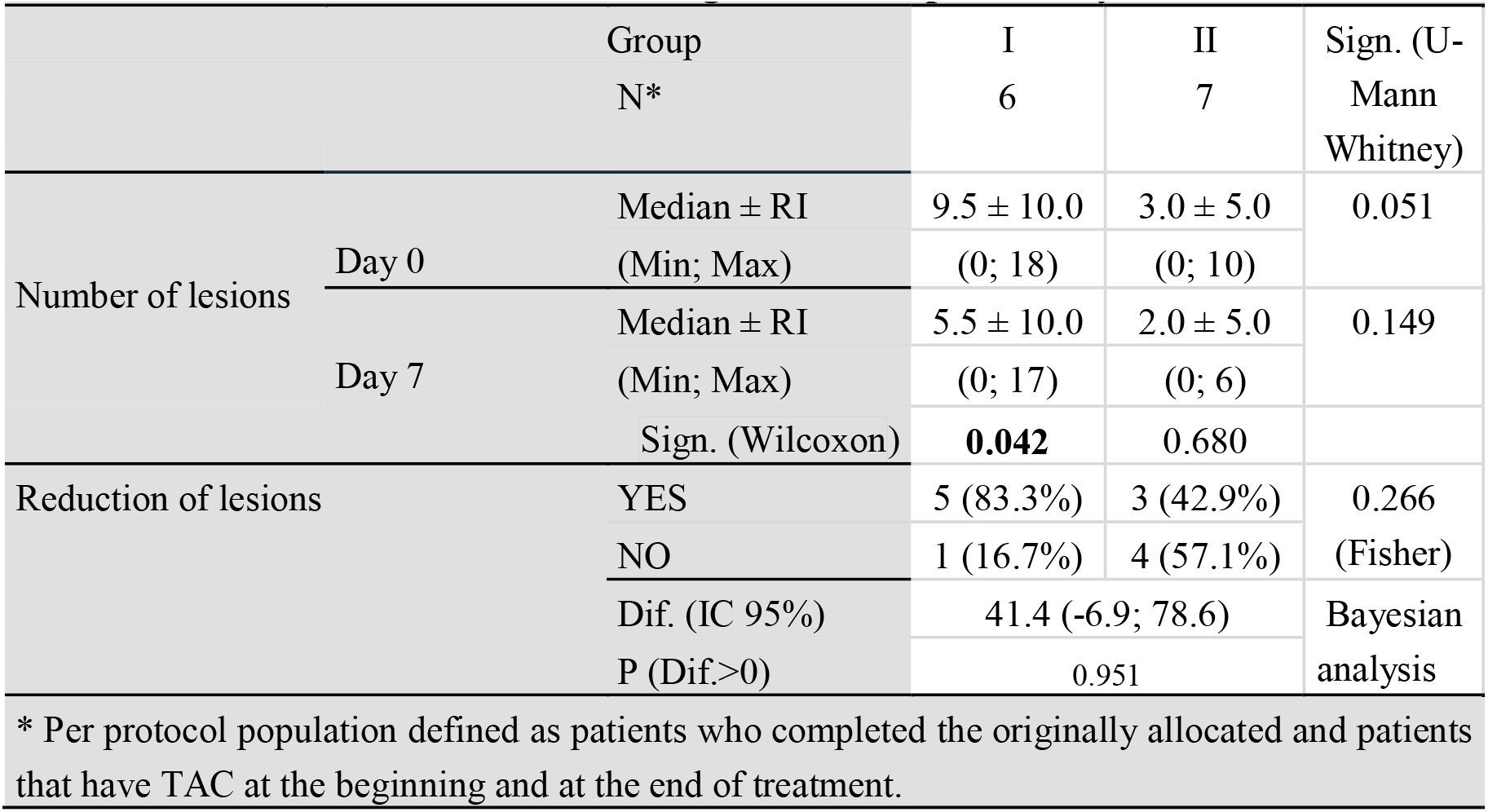
Chest-CT evolution considering number of pulmonary lesions.

The effect over lesion’s extent showed in Supp. 2 including those patients analyzed per protocol indicates that CIGB-325 treatment also reduced the median of lesion’s extent after consecutive-5 day regimen although not significantly. Otherwise, any kind of reduction was observed in the control group. Importantly, proportion of patients with reduction of lesion’s extent was higher in the CIGB-325 group (4/7) compared with control (1/9) (pDif > 0; 0.982) (Bayesian analysis).

The overall chest-CT response was also analyzed considering the lesion’s extent and the pattern of Covid-19 typical chest-CT abnormalities (Supp. 3). The analysis per protocol showed that 50% (3/6) of patients in the CIGB-325 group had a favorable overall chest-CT response compared with 28.6% (2/7) in the control (pDif > 0; 0.796) (Bayesian analysis). Importantly, any patient had progression in the CIGB-325 group and 28.6% (2/7) in the control group did it. Representative images of chest-CT evolution from one CIGB-325 treated patient are shown in Fig. 2.

**Figure 2:**
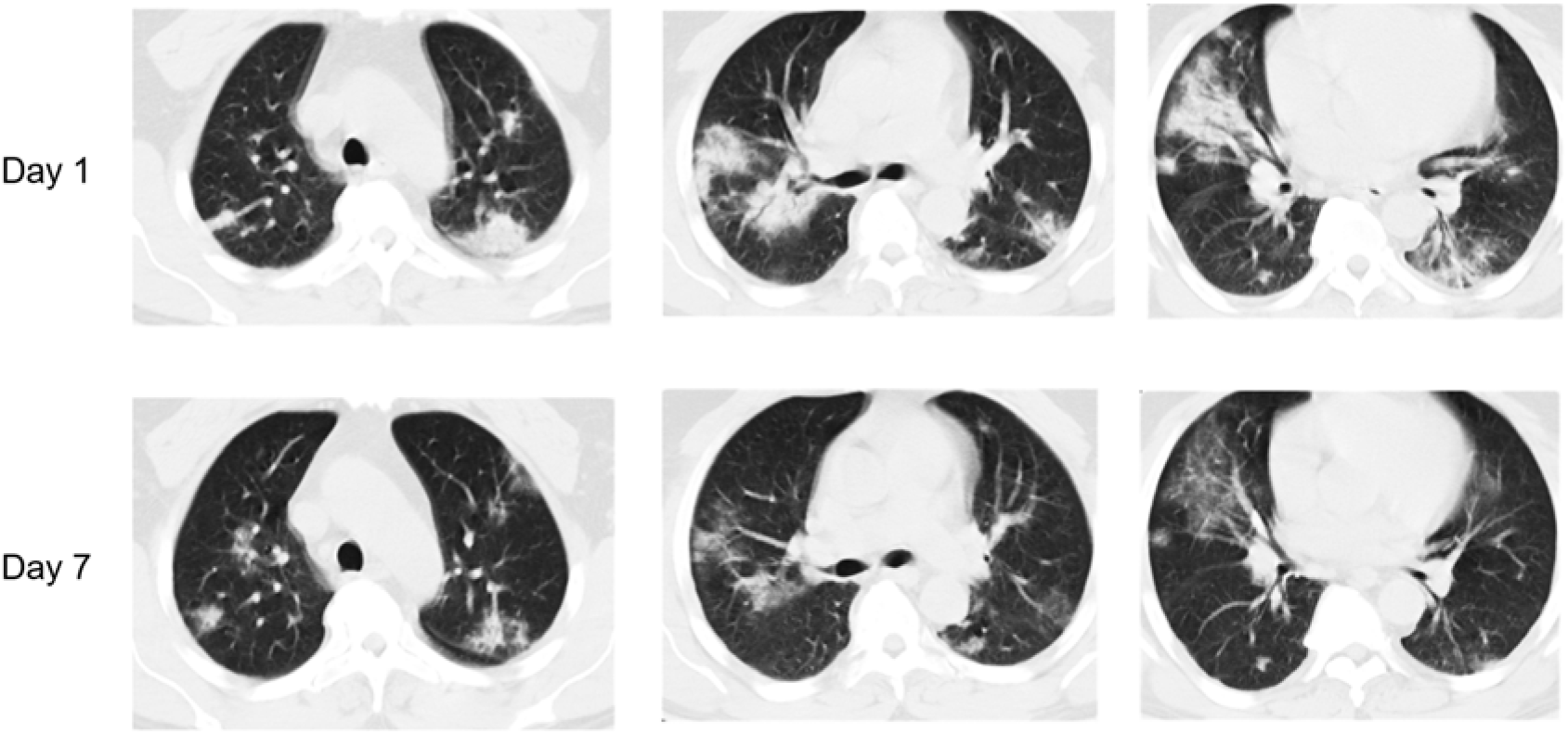
Chest-CT Evolution. Representative chest-CT images from one patient in the CIGB-325 group at day 0 and 7.

We also investigated the levels of Creatinine Phospho Kinase (CPK), Reactive C-Protein (RCP), and Lactate dehydrogenase (LDH) in plasma. Of note, data from Supp. 4 shows that CIGB-325 treatment significantly reduced the CPK (p = 0.007) and LDH (p = 0.028) serum levels. Also, the RCP values were lowered by CIGB-325 although with no statistical difference.

### Safety

Data from safety analysis indicated that intravenous CIGB-325 added to standard-of-care did increase both frequency of adverse events and patients with adverse events (Supp. 5). Particularly, pruritus, flushing and rash were increased by CIGB-325 treatment in the 100, 80 and 60 % of patients respectively. As previously observed during CIGB-325 intravenous administration, intensity of this kind of adverse events was mild and/or moderate in all of the patients.

## DISCUSSION

Protein kinase CK2 has been recently suggested as a relevant target to combat SARS-Cov2 infection because of its role on the viral particles egress once accumulated in filopodial protrusions possessing budding viral particles [19]. Although anti-SARS-Cov2 activity has been demonstrated through *in vitro* models by inhibiting protein kinase CK2, evidences of clinical benefit in Covid-19 for anti-CK2 approaches are not available at this.

CIGB-325 is an anti-CK2 synthetic peptide extensively assessed in cancer patients whose safety, tolerability and efficacy signs have been gathered from Phase I-II trials [22, 25]. This clinical trial investigated the short-term outcomes of intravenous CIGB-325 at 2.5 mg/kg in a consecutive-5 day regimen which was added to standard care used in Covid-19 disease in Cuba. Preliminary results suggest a quick clinical benefit with CIGB-325 treatment particularly evidenced by the chest-CT data at day 7 post-treatment initiation. Our trial found that this treatment regimen had clinical benefit for Covid-19 patients with lower respiratory tract involvement. Specifically, CIGB-325 treatment fostered reduction of both, the number pulmonary lesions, and lesion’s extent at day 7. Likewise, proportion of patients exhibiting this inhibitory effect is superior in the CIGB-325 group compared with standard care. When overall chest-CT response was analyzed by gathering number of lesions, lesion’s extent and the pattern of Covid-19 typical abnormalities, 50% of patients in the CIGB-325 group had favorable outcome compared with 20% in the standard-of-care. The odds of difference for the CIGB-325 treatment respect standard care were > 0.90 in all the chest-CT data by using a Bayesian model.

Recently, a clinical trial with RDV has reported clinical benefit in terms of the number of days to recovery of the hospitalized Covid-19 patients with lower respiratory tract involvement in 11 days (95% confidence interval [CI], 9 to 12) [15], however evidences of chest-CT improvement at day 7 were not provided.

The chest-CT response observed in our trial could be explained by direct antiviral activity of CIGB-325 in SARS-Cov2 infected pneumocytes [26]. Unpublished *in vitro* data from our lab have documented anti-SARS-Cov2 effect of CIGB-325 in Vero-E6 cells. Altogether, these findings further support an instrumental role of CK2 during SARS-Cov2 infection [19].

The improvement of chest-CT response by addition of CIGB-325 to standard-of-care could have a great clinical impact in avoiding progression to severity of Covid-19 patients. Additionally, our study shows that alpha 2b IFN + kaletra/hydroxychloroquine also reduces pulmonary lesions at day 7 although to a lesser extent. Such effect rather might be by the previously reported anti-SARS-Cov2 activity of type I IFNs when administered either by parenteral route or inhaled [27].

On the other hand, several laboratory parameters may facilitate the assessment of disease severity and rational triaging [28]. We found that CIGB-325 treatment decreased significantly the CPK levels compared with standard care which can suggest an early protective effect from tissue damage. Likewise, high serum CPK levels have been associated to rhabdomyolysis, weakness and heart injury in Covid-19 patients; therefore, these findings also support the clinical benefit of using CIGB-325 in this viral disease. Importantly, LDH and CRP serum levels which are considered inflammatory markers were also reduced after CIGB-325 treatment.

Improvement of the clinical status at day 7 also trended to be superior by adding CIGB-325 to the standard-of-care where 50% (4/8) patients experienced change towards a more favorable clinical category. Otherwise, standard-of-care alone improved clinical status in 25% (1/4).

The dynamic conversion from positive to negative of SARS-Cov2 RT-PCR at the nasopharyngeal swabs was very similar for both groups of treatments over the time. Addition of CIGB-325 to the standard care did not shorten the time to viral clearance respect to the standard care at least in the regimen tested in our clinical trial. Other protocols with higher frequency of CIGB-325 administration merit to be explored for that purpose in future trials. Nonetheless, the usefulness and feasibility of the viral end point in nasopharyngeal swabs to assess anti-SARS-Cov2 drugs has not been established yet. Instead, clinical endpoints seem to be more informative in the Covid-19 disease [11, 15, 24]

The safety profile of CIGB-325 plus standard-of-care was very similar to that observed for CIGB-325 alone in which a predominant pattern of transient histaminergic-like side effects has been observed [22]. However, intensity of the side effects was mild and/or moderate in all the patients and total resolution of them was achieved in no more than one hour after CIGB-325 administration. Thus, our study revealed that combination of intravenous CIGB-325 at 2.5 mg/kg with standard care for Covid-19 is a manageable and safe strategy to treat patients with this infectious disease.

In this exploratory study with a small sample size, randomization allowed a balance between the groups regarding the allocation of treatments but not regarding the prognostic variables. Also, although the sample size in our study is too small to permit generalizability, our data provide a window into what a larger trial might look like. Finally, our study gave interesting clues suggesting a quick clinical benefit at day 7 by using our anti-CK2 approach which has not been reported so far in Covid-19. Whether CIGB-325 prevents COVID-19 getting worse by inducing an antiviral effect in the lungs and preventing damage caused by the virus, is something that merit further investigation in larger clinical trials.

## CONCLUSION

Combination of CIGB-325 anti-CK2 peptide with alpha 2b-IFN/kaletra/hydroxychloroquine was adequately tolerated and superior in improving clinical and chest-CT response in Covid-19 patients as early as 7 days. SARS-Cov2 dynamics in nasopharyngeal swabs was similar for both treatments over the time. Our clinical findings support future studies to continue to improve patient outcomes with Covid-19 by optimizing CIGB-325 regimens and/or combining with other antiviral agents.

## Data Availability

All data referred in the manuscript is available.

## ACKNOWLEDGEMENTS

We thank Elizeth García, José Luis Rodriguez, Grettel Melo, Reinier Hernandez, Marel Alonso, Julio E Baldomero, and Francisco Hernandez from Clinical Department at the CIGB for their great contribution in the clinical trial execution. Also, we thank the Covid-19 diagnosis service from CIGB. This work was supported in part by the British Embassy in Havana with funds of the International Programme of the Foreign, Commonwealth and Development Office (FCDO).

## DISCLOSURES

The authors declare no conflict of interest.

## AUTHOR’S CONTRIBUTION

**Conceptualization:** IB, P AD, C.V, M.M.V, V.M, Y.P, S EP. **Methodology**: L.R.C, IB, P A D, M.M.V, R.S, A.G, D C, D.T., L.T, J.P, V.M., D.M.V, Y.V, R.R, M R, G.V.P., A.C.R, Y.P., SEP. **Software:** R.H, M.A. **Validation**: IB, C.V., G. P, A.G., R.R. **Formal analysis**: C.V. **Investigation:** L.R.C, A.R., R.S, A.G, D C, D.T, **Resources:** L.T, J.P, A.G.G, KG., S.G, M.E.F., J.F, Y.G, Y.A., R.P., L.P, R M., M.P., D.G., Y.O.T, D.P.M, E.A.D, Y.M., C A., Y.S., G.L. **Data curation**: I.E., M.G., S.M.M, R.H, M.A.. **Writing original draft:** SEP, C.V., IB. **Writing-Review and editing:** I B, Y.P, M R., L.D.R, P AD, C.V, S E P. **Visualization**: I B, C.V, Y.P, MR, SEP. **Supervision:** IB, PAD, M.M.V., V.M. **Project administration:** SEP, IB, G.G. **Funding acquisition:** G.G., SEP.

## APPENDIX

♣ATENEA-Co-300 group: Idelsis Esquivel, Maura García, Sara María Martínez, Ana Gloria Galarraga, Kenia Gómez, Sidalyn Gutiérrez, Maria E Fuerte, Jane Fernández, Yose Gamallo, Yanelis Alvarez, Yarelis Amador, Reynier Perez, Lisset Perurena, Rosa Martinez, Manuel Puerto, Daimays González, Yahima de la O Tamayo, Dianelis Pérez-Malo, Estela A Deyvis, Yuilyns Morales, Caridad Aguirre, Yuliet Sinclair, Reinier Hernández, Marel Alonso, Gilda Lemus.

## SUPPLEMENTARY MATERIALS

**Supplementary 1.** Demographic and Clinical Characteristics at Baseline.

|  |  |  |  |
| --- | --- | --- | --- |
| Characteristic | All<br>(N=20) | CIGB-325 +<br>Standard Care<br>(N=10) | Standard Care<br>(N=10) |
| **Age** (years) | 45.35±12.0 | 51.6±11.4 | 39.1±12.6 |
| **Sex No. (%)** Male | 14 (70) | 8 (80) | 6 (60) |
| Female | 6 (30) | 2 (20) | 4 (40) |
| **Skin color No. (%)** |  |  |  |
| White | 12 (60) | 5 (50) | 7 (70) |
| Mestizo | 2 (20) | 1 (10) | 1 (10) |
| Black | 6 (20) | 4 (40) | 2 (20) |
| **Coexisting conditions**<br>**No./Total No. (%)** |  |  |  |
| Hypertension | 5 (25) | 5 (50) | 0 (0.0) |
| Obesity | 5 (25) | 2 (20) | 3 (30) |
| Type 2 Diabetes | 0 (0.0) | 0 (0.0) | 0 (0.0) |
| Cancer | 1 (5) | 1 (10) | 0 (0.0) |
| Iron deficiency Anemia | 1 (5) | 0 (0.0) | 1 (10) |
| Hypothyroidism | 1 (5) | 0 (0.0) | 1 (10) |
| Glaucoma | 1 (5) | 0 (0.0) | 1 (10) |
| **Clinical status at**<br>**enrollment** |  |  |  |
| Asymptomatic | 14 (70) | 6 (60) | 8 (80) |
| Symptomatic | 6 (20) | 4 (40) | 2 (20) |
| **Clinical status at the**<br>**starting day of**<br>**treatment** |  |  |  |
| Asymptomatic | 9 (45) | 3 (30) | 6 (60) |
| Mild | 0 (0.0) | 0 (0.0) | 0 (0.0) |
| Moderate | 9 (45) | 5 (50) | 4 (40) |
| Severe | 2 (10) | 2 (20) | 0 (0.0) |
| **Symptoms** |  |  |  |
| Headache | 3 (15) | 2 (20) | 1 (10) |
| Anosmia | 1 (5) | 0 (0.0) | 1 (10) |
| Loss of taste | 1 (5) | 0 (0.0) | 1 (10) |
| Fever | 2 (10) | 2 (20) | 0 (0.0) |
| Dry cough | 2 (10) | 1 (10) | 1 (10) |
| Loss of appetite | 1 (5) | 1 (10) | 0 (0.0) |
| Shortness of breath | 1 (5) | 0 (0.0) | 1 (10) |
| Diarrhea | 1 (5) | 1 (10) | 0 (0.0) |

**Supplementary 2:**
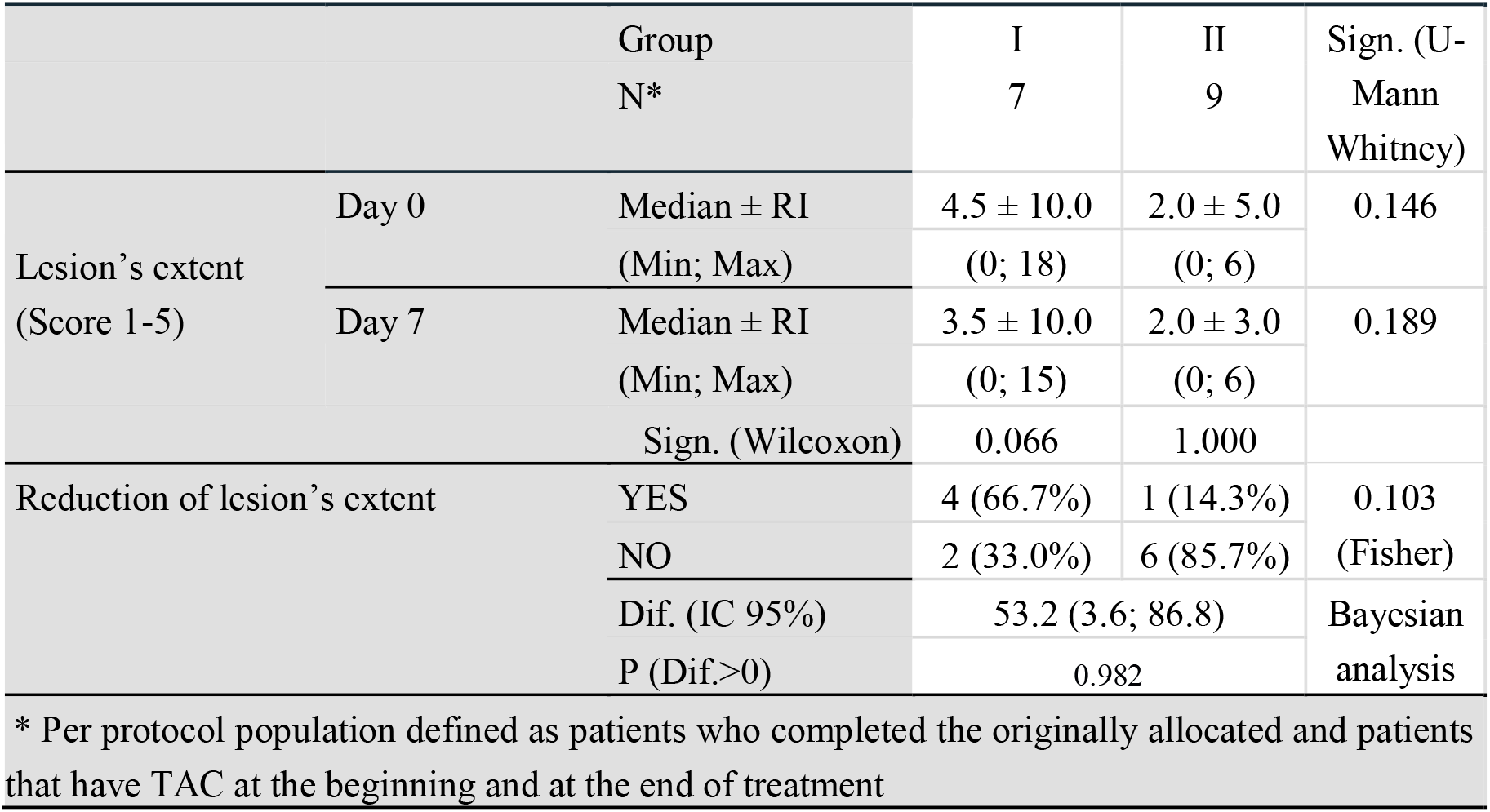
Chest-CT evolution considering lesion’s extent.

**Supplementary 3:**
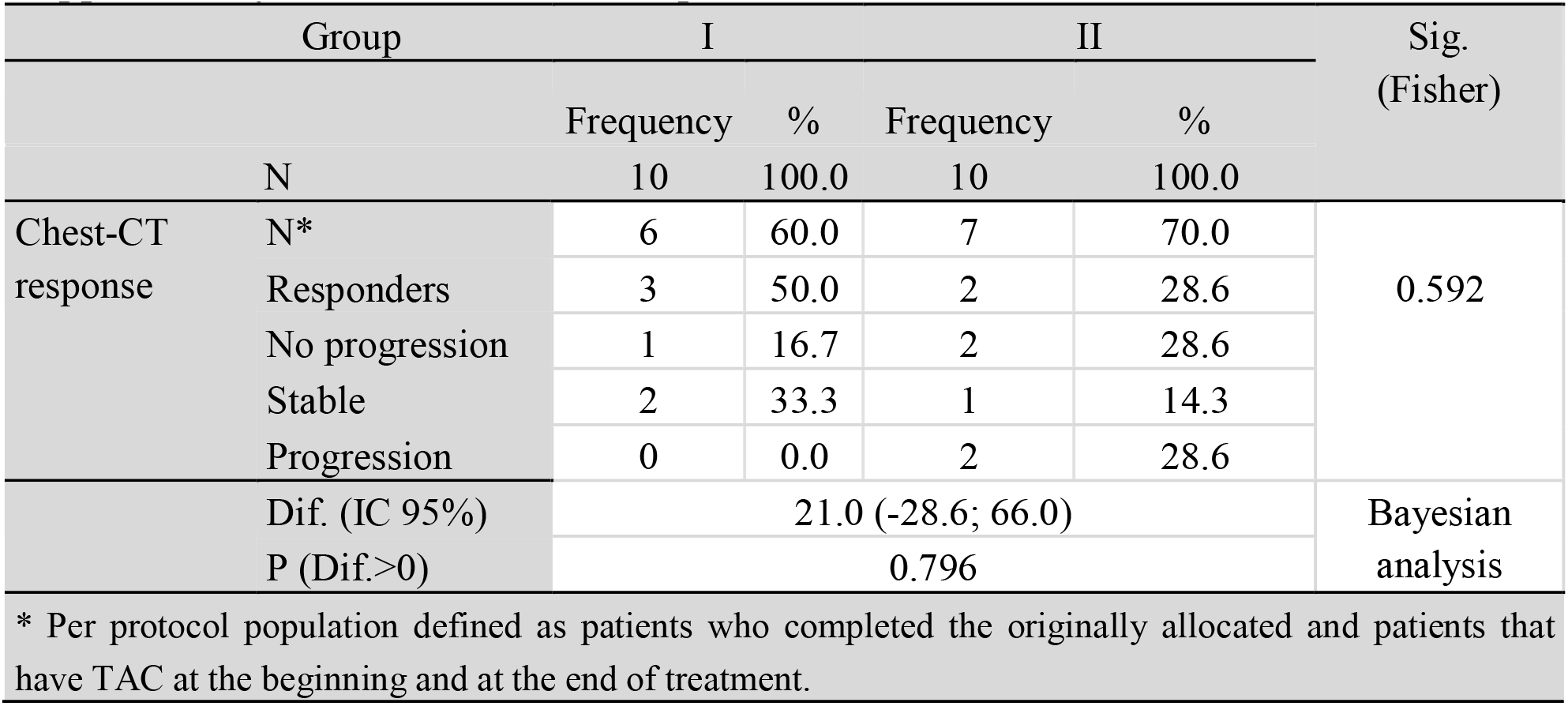
Overall chest-CT response.

**Supplementary 4:**
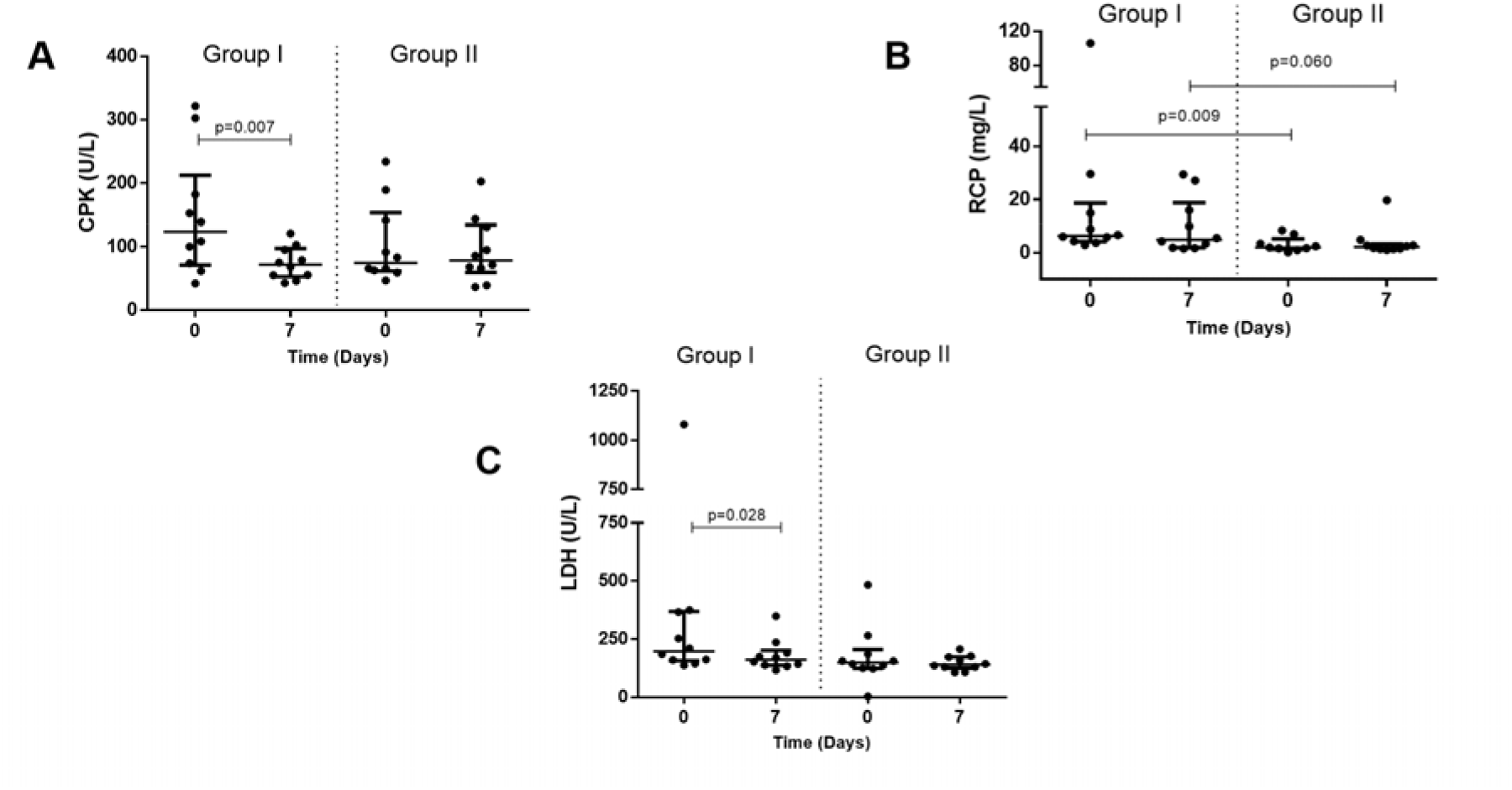
Serum levels of Covid-19 linked markers. Serum levels of CPK (**A**), RCP (**B**) and LDH (**C**). Group I: CIGB-325 + Standard-of-care. Group II: Standard-of-care.

**Supplementary 5:** Adverse events.

|  | Group I |  |  |  | Group II |  |  |  |
| --- | --- | --- | --- | --- | --- | --- | --- | --- |
|  | Patients |  | Events |  | Patients |  | Events |  |
|  | Frequency | % | Frequency | % | Frequency | % | Frequency | % |
| N | 10 | 100 | 109 | 100 | 10 | 100 | 7 | 100 |
| Pruritus | 10 | 100.0 | 48 | 44.0 | -- | -- | -- | -- |
| Rash | 6 | 60.0 | 12 | 11.0 | -- | -- | -- | -- |
| Flush | 8 | 80.0 | 23 | 21.1 | -- | -- | -- | -- |
| Hot flashes | 5 | 50.0 | 12 | 11.0 | -- | -- | -- | -- |
| Wheals | 3 | 30.0 | 5 | 4.6 | -- | -- | -- | -- |
| Bradycardia | 1 | 10.0 | 1 | 0.9 | -- | -- | -- | -- |
| Cramps | 2 | 20.0 | 2 | 1.8 | 1 | 10.0 | 1 | 14.3 |
| Trembling | 2 | 20.0 | 2 | 1.8 | -- | -- | -- | -- |
| Chills | 1 | 10.0 | 1 | 0.9 | -- | -- | -- | -- |
| Nausea | 1 | 10.0 | 1 | 0.9 | 1 | 10.0 | 2 | 28.6 |
| Tinnitus | 1 | 10.0 | 2 | 1.8 | -- | -- | -- | -- |
| Diarrhea | -- | -- | -- | -- | 1 | 10.0 | 2 | 28.6 |
| Headache | -- | -- | -- | -- | 1 | 10.0 | 2 | 28.6 |
| Group I: CIGB-325 plus standard-of-care |  |  |  | Group II: Standard-of-care |  |  |  |  |

## Notes

### Competing Interest Statement

The authors have declared no competing interest.

### Clinical Trial

IG/CIGB300I/CV/2001

### Author Declarations

The Ethics Committee for Clinical Research in the Luis Diaz Soto Hospital and Cuban Regulatory Agency (CECMED) approved the trial. The study complied with the Good Clinical Practices and the precepts established in the Declaration of the Helsinki World Medical Association. All patients met the inclusion criteria described in the protocol and signed the informed consent.

## References

1. https://www.who.int/docs/default-source/coronaviruse/situation-reports/20200809-covid-19-sitrep-202.

2. https://coronavirus.jhu.edu/map.html.

3. Cohen, P. A.; Hall, L.; John, J. N.; Rapoport, A. B. In The early natural history of SARS-CoV-2 infection: Clinical observations from an urban, ambulatory COVID-19 clinic, Mayo Clinic Proceedings, 2020; Elsevier: 2020; pp 1124–1126.

4. Wu, Z.; McGoogan, J., Outbreak in China: Summary of a Report of 72314 Cases from the Chinese Center for Disease Control and Prevention. JAMA 2020, 10.

5. Wu, J.; Mafham, M.; Mamas, M.; Rashid, M.; Kontopantelis, E.; Deanfield, J.; de Belder, M.; Gale, C. P., Place and underlying cause of death during the COVID19 pandemic: retrospective cohort study of 3.5 million deaths in England and Wales, 2014 to 2020. *medRxiv* 2020.

6. Phillips, R. O.; Robert, J.; Abass, K. M.; Thompson, W.; Sarfo, F. S.; Wilson, T.; Sarpong, G.; Gateau, T.; Chauty, A.; Omollo, R., Rifampicin and clarithromycin (extended release) versus rifampicin and streptomycin for limited Buruli ulcer lesions: a randomised, open-label, noninferiority phase 3 trial. The Lancet 2020.

7. Pushpakom, S.; Iorio, F.; Eyers, P. A.; Escott, K. J.; Hopper, S.; Wells, A.; Doig, A.; Guilliams, T.; Latimer, J.; McNamee, C., Drug repurposing: progress, challenges and recommendations. Nature reviews Drug discovery 2019, 18, (1), 41–58.

8. Ohashi, H.; Watashi, K.; Saso, W.; Shionoya, K.; Iwanami, S.; Hirokawa, T.; Shirai, T.; Kanaya, S.; Ito, Y.; Kim, K. S., Multidrug treatment with nelfinavir and cepharanthine against COVID-19. *bioRxiv* 2020.

9. Bao, L.; Deng, W.; Gao, H.; Xiao, C.; Liu, J.; Xue, J.; Lv, Q.; Liu, J.; Yu, P.; Xu, Y., Lack of reinfection in rhesus macaques infected with SARS-CoV-2. *bioRxiv* 2020.

10. Cao, B.; Wang, Y.; Wen, D.; Liu, W.; Wang, J.; Fan, G.; Ruan, L.; Song, B.; Cai, Y.; Wei, M., A trial of lopinavir-ritonavir in adults hospitalized with severe Covid-19. New England Journal of Medicine 2020.

11. Wang, Y.; Zhang, D.; Du, G.; Du, R.; Zhao, J.; Jin, Y.; Fu, S.; Gao, L.; Cheng, Z.; Lu, Q., Remdesivir in adults with severe COVID-19: a randomised, double-blind, placebo-controlled, multicentre trial. The Lancet 2020.

12. Borba, M. G. S.; Val, F. F. A.; Sampaio, V. S.; Alexandre, M. A. A.; Melo, G. C.; Brito, M.; Mourao, M. P. G.; Brito-Sousa, J. D.; Baía-da-Silva, D.; Guerra, M. V. F., Effect of high vs low doses of chloroquine diphosphate as adjunctive therapy for patients hospitalized with severe acute respiratory syndrome coronavirus 2 (SARS-CoV-2) infection: a randomized clinical trial. JAMA network open 2020, 3, (4), e208857–e208857.

13. Cai, Q.; Yang, M.; Liu, D.; Chen, J.; Shu, D.; Xia, J.; Liao, X.; Gu, Y.; Cai, Q.; Yang, Y., Experimental treatment with favipiravir for COVID-19: an open-label control study. Engineering 2020.

14. Bauchner, H.; Fontanarosa, P. B., Randomized clinical trials and COVID-19: managing expectations. Jama 2020.

15. Beigel, J. H.; Tomashek, K. M.; Dodd, L. E.; Mehta, A. K.; Zingman, B. S.; Kalil, A. C.; Hohmann, E.; Chu, H. Y.; Luetkemeyer, A.; Kline, S., Remdesivir for the treatment of Covid-19—preliminary report. New England Journal of Medicine 2020.

16. Ahidjo, B. A.; Loe, M.; Ng, Y. L.; Mok, C. K.; Chu, J. J. H., Current Perspective of Antiviral Strategies against COVID-19. ACS Infectious Diseases 2020.

17. Meggio, F.; Pinna, L. A., One-thousand-and-one substrates of protein kinase CK2? FASEB jtournal: tofficial publicatiton tof the Federatiton tof American Stocieties ftor Experimental Bitoltogy 2003, 17, (3), 349–68.

18. Keating, J. A.; Striker, R., Phosphorylation events during viral infections provide potential therapeutic targets. Reviews in medical virology 2012, 22, (3), 166–81.

19. Bouhaddou, M.; Memon, D.; Meyer, B.; White, K. M.; Rezelj, V. V.; Marrero, M. C.; Polacco, B. J.; Melnyk, J. E.; Ulferts, S.; Kaake, R. M., The global phosphorylation landscape of SARS-CoV- 2 infection. Cell 2020, 182, (3), 685–712. e19.

20. Perea, S. E.; Reyes, O.; Puchades, Y.; Mendoza, O.; Vispo, N. S.; Torrens, I.; Santos, A.; Silva, R.; Acevedo, B.; López, E.; Falcón, V.; Alonso, D. F., Antitumor effect of a novel proapoptotic peptide that impairs the phosphorylation by the protein kinase 2 (casein kinase 2). Cancer Res 2004, 64, (19), 7127–9.

21. Perera, Y.; Ramos, Y.; Padrón, G.; Caballero, E.; Guirola, O.; Caligiuri, L. G.; Lorenzo, N.; Gottardo, F.; Farina, H. G.; Filhol, O., CIGB-300 anti-cancer peptide regulates the protein kinase CK2-dependent phosphoproteome. Mol Cell Biochem 2020.

22. García-Diegues, R.; de la Torre-Santos, A., Phase I Study of CIGB-300 Administered Intravenously in Patients with Relapsed/Refractory Solid Tumors. ARCHIVOS DE MEDICINA 2018, 1, (1), 4.

23. Fernández Águila, J. D.; García Vega, Y.; Ríos Jiménez, R. O.; López Sacerio, A.; Rodríguez Rodríguez, C. R.; Rodríguez Fraga, Y.; Valenzuela Silva, C., Safety of intravenous application of cigb-300 in patients with hematological malignancies. EHPMA study. Revista Cubana de Hematología, Inmunología y Hemoterapia 2016, 32, (2), 236–248.

24. https://www.fda.gov/drugs/guidance-compliance-regulatory-information/guidances-drugs.

25. Perea, S. E.; Baladrón, I.; Valenzuela, C.; Perera, Y., CIGB-300: A peptide-based drug that impairs the Protein Kinase CK2-mediated phosphorylation. Semin Oncol2018, 45, (1-2), 58–67.

26. Carsana, L.; Sonzogni, A.; Nasr, A.; Rossi, R. S.; Pellegrinelli, A.; Zerbi, P.; Rech, R.; Colombo, R.; Antinori, S.; Corbellino, M., Pulmonary post-mortem findings in a series of COVID-19 cases from northern Italy: a two-centre descriptive study. The Lancet Infectious Diseases 2020.

27. Pereda, R.; Gonzalez, D.; Rivero, H. B.; Rivero, J. C.; del Rosario Lopez, L.; Mezquia, N.; Venegas, R.; Betancourt, J. R.; Dominguez, R. E., Therapeutic effectiveness of interferon alpha 2b treatment for COVID-19 patient recovery. *medRxiv* 2020.

28. Velavan, T. P.; Meyer, C. G., Mild versus severe COVID-19: Laboratory markers. International Journal of Infectious Diseases 2020, 95, 304–307.

